# Adults prenatally exposed to the Dutch Famine exhibit a metabolic signature associated with a broad spectrum of common diseases

**DOI:** 10.1101/2024.04.04.24305284

**Authors:** M. Jazmin Taeubert, Thomas B. Kuipers, Jiayi Zhou, Chihua Li, Shuang Wang, Tian Wang, Elmar W. Tobi, BBMRI-NL Metabolomics consortium, Daniel W. Belsky, L. H. Lumey, Bastiaan T. Heijmans

## Abstract

**Background:** Exposure to famine in the prenatal period is associated with an increased risk of metabolic disease, including obesity and type-2 diabetes. We employed nuclear magnetic resonance (NMR) metabolomic profiling to provide a deeper insight into the metabolic changes associated with survival of prenatal famine exposure during the Dutch Famine at the end of World War II and explore their link to disease.

**Methods:** NMR metabolomics data were generated from serum in 480 individuals prenatally exposed to famine (mean 58.8 years, 0.5 SD) and 464 controls (mean 57.9 years, 5.4 SD). We tested associations of prenatal famine exposure with levels of 168 individual metabolic biomarkers and compared the metabolic biomarker signature of famine exposure with those of 154 common diseases.

**Results:** Prenatal famine exposure was associated with higher concentrations of branched-chain amino acids ((iso)-leucine), aromatic amino acid (tyrosine), and glucose in later life (0.2-0.3 SD, p < 3×10^-3^). The metabolic biomarker signature of prenatal famine exposure was positively correlated to that of incident type-2 diabetes (r = 0.77, p = 3×10^-27^), also when re-estimating the signature of prenatal famine exposure among individuals without diabetes (r = 0.67, p = 1×10^-18^). Remarkably, this association extended to 115 common diseases for which signatures were available (0.3 :< r :< 0.9, p < 3.2×10^-4^). Correlations among metabolic signatures of famine exposure and disease outcomes were attenuated when the famine signature was adjusted for body mass index.

**Conclusions:** Prenatal famine exposure is associated with a metabolic biomarker signature that strongly resembles signatures of a diverse set of diseases, an observation that can in part be attributed to a shared involvement of obesity.

## Background

Metabolomics is a powerful tool for illuminating molecular phenotypes underpinning disease (1). With nuclear magnetic resonance (NMR) approaches, metabolomics is now possible within large-scale epidemiologic studies and biobanks. Within these settings, NMR metabolomics is revealing a range of common and unique features to a broad spectrum of diseases (2–5). Less is known about how the metabolome may reflect or mediate effects of prenatal exposure histories on disease pathogenesis. Here, we investigate the long-term metabolomic sequelae of gestational exposure to famine, an established risk factor for the development of metabolic disease (6,7).

The Dutch Hunger Winter of 1944-1945, a 6-month famine at the end of World War II, provides a unique setting to study the long-term effects of an adverse prenatal environment (6,8,9). Previous studies revealed that prenatal famine exposure is associated with an increased risk in unfavourable metabolic phenotypes in adulthood including increased fasting glucose and triglyceride levels, obesity, and type-2 diabetes (10–17). These associations have also been observed for other historical famines (6). To date, a comprehensive view of metabolic changes linked to prenatal famine exposure is lacking. In this study, we seek to provide a deeper insight into the metabolomic profile associated with prenatal famine exposure.

We profiled samples for 944 participants from the Dutch Hunger Winter Families Study using nuclear magnetic resonance (NMR) metabolomics. We compared prenatal famine-exposed individuals to unexposed control participants on 168 different serum metabolic biomarkers and also compared the metabolome-wide signature of prenatal famine exposure to an atlas of signatures marking risk of a range of common diseases in order to characterize the phenotype of prenatal famine exposure. Our study reveals specific metabolic biomarker alterations and broader connections with a range of chronic diseases, providing new insights into how prenatal famine exposure shapes health across the life course.

## Methods

### Study population

The Dutch Hunger Winter Families study (DHWFS) is described in detail elsewhere (18). In short, historical birth records were retrieved from three institutions in famine-exposed cities of all singleton births between 1 February 1945 and 31 March 1946 and a systematic sample of births born in 1943 or 1947. From these records we identified infants whose mothers were exposed to the famine during or immediately preceding that pregnancy and unexposed time-controls born before or after the famine. These individuals were invited to participate in a telephone interview and in a clinical examination, together with a same-sex sibling not exposed to the famine (family-control).

The Dutch Hunger Winter Families study was approved by the Medical Ethics Committee of Leiden University Medical Center (P02.082) and the participants provided verbal consent at the start of the telephone interview and written informed consent at the start of the clinical examination.

We conducted 1,075 interviews and 971 clinical examinations between 2003 and 2005. One non-biological sibling identified with genetic analyses was excluded from the cohort. NMR metabolomics profiling was performed on serum samples of 962 individuals. Our sample for this study included 944 individuals after excluding non-fasted samples (n = 17) and an outlier in the metabolomics dataset as identified with principal component analysis (n = 1) (**Supplemental Fig. 1**).

### Famine exposure definitions

Food rations were distributed centrally and below 900 kcal/day between November 26, 1944 and May 15, 1945 (8). We defined famine exposure by the number of weeks during which the mother was exposed to < 900 kcal/day after the last menstrual period (LMP) recorded on the birth record (18). For analysis of timing of gestational exposure, we subdivide the gestational period into units of 10 weeks. We considered the mother exposed in gestational weeks 1-10, 11-20, 21-30, or 31 to delivery if these gestational time windows were entirely contained within this period and had an average exposure of < 900kcal/day during an entire gestation period of 10 weeks. As the famine lasted 6-months some participants were exposed to famine during two adjacent 10-week periods. In chronological order, pregnancies with LMP between 30 April 1944 and 24 August 1944 were considered exposed in weeks 31 to delivery; between 9 July 1944 and 15 October 1944 in pregnancy weeks 21-30; between 17 September 1944 and 24 December 1944 in pregnancy weeks 11-20, between 26 November 1944 and 4 March 1945 in pregnancy weeks 1-10. Individuals with a LMP between 4 February and 12 May 1945 were exposed to an average of < 900kcal/day for less than 10 weeks before conception and up to 8 weeks post-conception, are denoted as the weeks 9-0 weeks group. We defined individuals exposed to one or at most two of these definitions exposed to ‘any’ gestational exposure.

### Characteristics

Information on health history, including information on the use of cholesterol-lowering drugs, was collected through telephone interviews. Measurement of height was carried out to the nearest millimeter using a portable stadiometer (Seca), and body weight was measured to the nearest 100 g by a portable scale (Seca). BMI was calculated from these measures (weight (kg) / [height (m)]^2^). Cholesterol measures were reported previously (14) and were assessed using standard enzymatic assays. LDL cholesterol was calculated for individuals with a triglyceride concentration lower than 400 mg/dl using the Friedewald formula. A blood draw was performed at the start of a 75-g oral glucose test, and fasted glucose was quickly assessed in serum by hexokinase reaction on a Modular P800 (Roche). The presence of type-2 diabetes was either determined through previous health history or defined as fasting glucose ≥ 7.0mmol/l or 2hr glucose tolerance test ≥ 11.1mmol/l (19).

### Metabolic biomarker quantification

Metabolic biomarkers were measured from serum samples using a high-throughput ^1^H-NMR metabolomics platform developed by Nightingale Health Ltd. (Helsinki, Finland; nightingalehealth.com; biomarker quantification version 2021). Details of the procedure and application of the NMR metabolomics platform have been described elsewhere (20,21). This method provides simultaneous quantification of 168 directly measured and 81 derived metabolic biomarkers, including 37 clinically validated metabolic biomarkers certified for diagnostics use. The metabolic biomarkers measured include amino acids, ketone bodies, lipids, fatty acids, and lipoprotein subclass distribution, particle size and composition. A subset of the biomarkers was selected for inclusion in the presented analysis, focusing on the 168 directly measured metabolic biomarkers.

Missing values were set to the minimum value for each metabolic measure. A value of one was added to all metabolic biomarkers containing zeroes (i.e. *x* + 1), which indicated that they were below the limit of quantification. All metabolic biomarkers were then natural logarithmic transformed to obtain an approximately normal distribution. The metabolic biomarkers were subsequently scaled to standard deviation (SD) units (mean 0, SD 1) for use in the analysis.

### Genotype data generation and polygenic scores

From our metabolomics sample population, 931 individuals also had genotype data available. Genotype data were measured using the Illumina Infinium^TM^ Global Screening Array (GSA) genotyping platform (version 24 v3.0. Illumina Inc., San Diego, USA) by the Human Genomics Facility in the Genetic Laboratory Rotterdam (Rotterdam, the Netherlands). Imputation was performed using the 1000G P3v5 reference panel (22). Polygenic scores were calculated for tyrosine, leucine and glucose levels with the PRSice-2 software using the independent hits of publicly available genome-wide association study (GWAS) summary statistics (**Supplemental Table 3**) (23). The base GWAS study utilized the same NMR platform and had participants with the same ancestry (European) as those in our study (24). The polygenic scores were residualized on the first ten genetic principal components and subsequently scaled to standard deviation (SD) units (mean 0, SD 1) for analysis.

### Statistical analysis

All analyses were performed in the R programming environment (R version 4.2.2).

For all linear regression analyses, we used linear regression within a generalized estimating equations framework to account for the correlation between sibships (R geepack package, version 1.3.9) (25) and adjusted for age, sex and cholesterol-lowering medication.

We first validated the NMR measurements by testing the consistency between glucose, triglycerides, total cholesterol, LDL, and HDL cholesterol measured by routine clinical chemistry and Nightingale Health NMR (**Supplemental Fig. 2**). Consistent with previous studies, correlations were high (r ≥ 0.9) for all metabolic biomarkers tested (26,27). We subsequently tested whether previously observed associations between prenatal famine exposure and these five metabolic biomarkers as measured by routine clinical chemistry were consistently found when the same biomarkers were measured using Nightingale Health NMR. Next, we performed a metabolome-wide association study of prenatal famine exposure by assessing the relationship between famine exposure and 168 metabolic biomarkers. Due to the correlated nature of the metabolic biomarkers, 95% of the variation in the 168 metabolic biomarkers was explained by 14 principal components. Therefore, as previously described (28–30), we corrected for 14 independent tests using Bonferroni multiple testing correction (p value = 0.05/14 = 3.57×10^−3^). Sensitivity analyses were performed to assess the robustness of the results of the metabolome-wide association study of prenatal famine. First, to assess the effect of famine exposure independent of BMI or type-2 diabetes, the main model was additionally adjusted for BMI and type-2 diabetes. Second, to determine the extent to which the observed associations between prenatal famine exposure and metabolic biomarkers levels could be attributed to genetics, the main model was additionally adjusted for the polygenic scores of the metabolic biomarkers. Third, to check for potential differences between sexes, sex-stratified analyses were performed adjusting for the same covariates as the main model and an interaction term for sex and metabolic marker was included in the model to test whether potential differences were statistically significant. Fourth, potential gestation timing specific effects of famine exposure were examined by subdividing famine exposure into 5 gestational time windows. In the regression analysis, the single indicator of famine exposure was replaced with indicator variables identifying exposure within each of the gestational time windows.

To expand our analysis from focusing on individual metabolic biomarkers to broader metabolic biomarker signatures associated with disease, we compared the metabolic biomarker signature associated with prenatal famine exposure to the metabolic biomarker signature predicting the future risk of type-2 diabetes. For this we utilized published results from a metabolome-wide study on incident type-2 diabetes using UK Biobank data (2). Specifically, we correlated the effect sizes of famine exposure in DHWFS with the effect sizes in the UK Biobank for incident type-2 diabetes across the 135 shared metabolic biomarkers in both datasets.

We then extended our analysis and correlated the metabolic profile of prenatal famine to the publicly availably metabolic signatures of a large set of diseases as estimated with UK Biobank data (4). Out of 674 incident diseases available in the metabolomics atlas, we selected those with at least 1000 cases (out of a total population ranging from ∼103,300 to ∼118,000) to represent common diseases (n = 162), and those with at least one significant association with a metabolic biomarker in the UK Biobank analysis (p < 5×10^-4^) (n = 154), resulting in 154 diseases. The effect sizes of the overlapping 168 metabolic biomarkers were correlated between prenatal famine and each disease. We corrected for multiple testing using Bonferroni correction (p value = 0.05/154 = 3.2×10^-4^). Finally, we re-estimated the effect sizes of the association between prenatal famine exposure and all 168 metabolic biomarkers, while additionally adjusting for BMI. We then repeated the correlation analysis comparing this BMI-adjusted prenatal famine metabolic biomarker signature with the 154 metabolic biomarker disease signatures from the UK Biobank.

## Results

### Population characteristics

Within the Dutch Hunger Winter Families Study, fasting NMR metabolomics data were available for 944 study participants. Among these participants, 480 (51%) were prenatally exposed to famine, and 464 (49%) were controls (including unexposed time controls born at the same institution as the exposed individuals and unexposed same-sex sibling controls both born either before or conceived after the famine). As previously reported, famine-exposed participants had an increased BMI (11) and a higher prevalence of type-2 diabetes (16) and controls were on average 0.9 years younger than famine-exposed. No differences were observed in sex or the use of cholesterol lowering medication (**Table 1**).

**Table 1.** Population characteristics.

|  | **Controls** | **Famine-exposed** | **p value** |
| --- | --- | --- | --- |
|  | **(*n* = 464)** | **(*n* = 480)** |  |
| Age, years (SD) | 57.9 (5.4) | 58.8 (0.5) | 1.1x10<sup>-3</sup> |
| Sex, males, n (%) | 200 (43.1) | 225 (46.9) | 0.24 |
| Use of cholesterol-lowering medication, n (%) | 55 (11.9) | 61 (12.7) | 0.69 |
| Body mass index, kg/m<sup>2</sup> (SD) | 27.0 (4.2) | 28.2 (4.8) | 1.6x10<sup>-4</sup> |
| Type-2 diabetes, n (%) | 38 (8.2) | 61 (12.8) | 0.02 |

Values are means (standard deviation) or numbers of subjects (valid %) shown for famine-exposed and controls of the study population. Comparing the two categories by a two-sample t-test or chi-square test, as appropriate.

### Validation of NMR metabolomics measures

We first sought to validate the newly measured metabolomics measures in our study by correlating them with previously measured clinical chemistry data available for five metabolic biomarkers, namely fasted glucose, triglycerides, total cholesterol, LDL, and HDL cholesterol (14,17). The correlations were all high (r ≥ 0.9) in line with previous studies (**Supplemental Fig. 2**) (4,31). Next, we examined the associations between prenatal famine exposure and these five metabolic biomarkers, as measured by clinical chemistry or NMR, and found the effect sizes to be consistent between the two measurements (**Supplemental Table 1)**.

### Metabolome-wide association study on prenatal famine exposure

Next, we examined the association of any prenatal famine exposure with all 168 metabolic biomarkers individually. Prenatal famine exposure was associated with higher tyrosine (effect size 0.28 SD), leucine (0.21 SD), glucose (0.23 SD), and isoleucine (0.18 SD) concentrations (p<3×10^-3^; all analyses adjusted for age, sex, and use of cholesterol-lowering drugs) (**Fig. 1A, Supplemental Table 2**).

**Fig 1.**
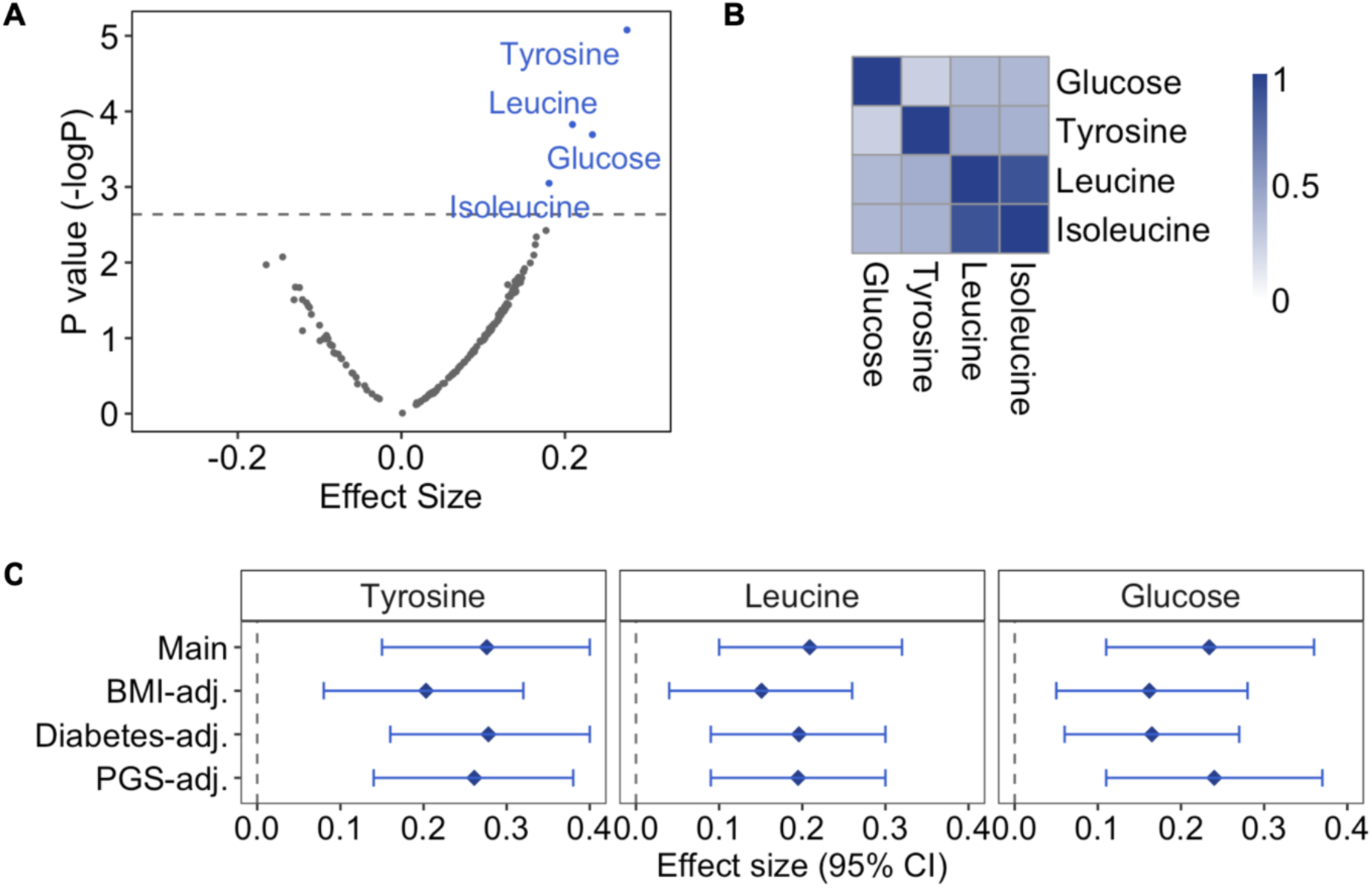
Metabolome-wide association study on prenatal famine exposure. **A.** Association of prenatal famine exposure with 168 metabolic biomarkers. Regression models were adjusted for age, sex and cholesterol-lowering medication and correlation within sibships were controlled for (main model). Scattered points represent metabolic biomarkers: the x-axis shows the effect size for the association of famine with the respective metabolic biomarker, while the y-axis is negative log of the p value. The grey line represents the significance threshold for this analysis (p value = 3.57×10^−3^). **B.** Heatmap showing the correlation of famine-associated metabolic biomarkers. Pearson’s correlation was calculated for each metabolic biomarker pair. **C.** Sensitivity analyses on famine-associated metabolic biomarkers. Main: main model; BMI-adjusted: main model additionally adjusting for BMI; Diabetes-adjusted: main model additionally adjusted for type-2 diabetes; polygenic score (PGS)-adjusted: main model additionally adjusted for the polygenic score of the metabolic biomarkers. Effect estimates and 95% confidence intervals are depicted for each model and are reported in standard-deviation (SD) units of the log-transformed metabolic biomarkers.

We performed three sets of follow-up analyses for tyrosine, leucine, and glucose to gain further insight into these associations. Isoleucine was excluded because it was highly correlated with leucine (r = 0.9), both are branched-chain amino acids, while leucine showed the stronger association with famine exposure (**Fig. 1B, Supplemental Fig. 3**). First, the associations between prenatal famine exposure and tyrosine, leucine, and glucose remained after including BMI or type-2 diabetes as a covariate in the model (**Fig. 1C**). Second, we also considered genetics as a potential explanatory factor for these associations. Since the polygenic scores of each metabolic biomarker explained only approximately 1%-5% of their variance (**Supplemental Table 3**), the associations were not affected by including the polygenic scores as covariates (**Fig. 1C**). Third, we explored whether associations between famine exposure and the three metabolic biomarkers were dependent on sex or the timing of exposure during gestation (**Supplemental Fig. 4**). Effect sizes across different exposure timing subgroups were similar to our estimates of the main analysis. In the sex stratified analysis, the effect sizes for glucose and tyrosine were lower in females than males, but we found no statistical evidence for effect modification (interaction p values > 0.39).

### Comparison to metabolic biomarker signatures of diseases

The individual metabolic biomarkers associated with prenatal famine exposure were previously linked to type-2 diabetes (32). To further investigate whether these associations reflect an increased risk of type-2 diabetes among individuals prenatally exposed to famine, we utilized a previously reported metabolic biomarker signature of incident type-2 diabetes from UK Biobank (2) and compared it to the complete set of biomarker associations in our study. Specifically, we took the effect sizes of 135 metabolic biomarkers for the risk of type-2 diabetes and compared them to the effect sizes we observed for prenatal famine. The metabolic biomarker signature of prenatal famine exposure was highly correlated with that of incident type-2 diabetes (r = 0.77, p = 3×10^-27^; **Fig. 2A**). This similarity persisted when we re-estimated the effect sizes for prenatal famine exposure after excluding participants with type-2 diabetes (r = 0.67, p = 1×10^-18^) (**Fig. 2B**).

**Fig 2.**
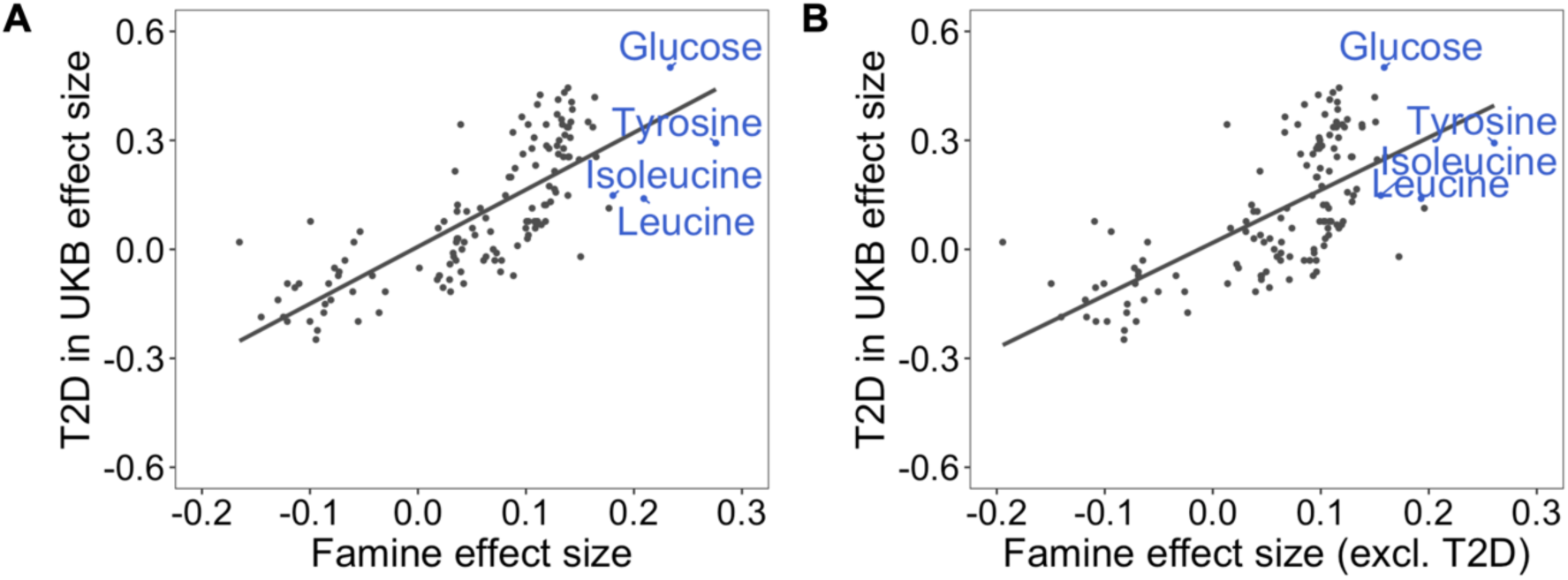
Correlation analysis of the metabolic biomarker signature associated with famine and the metabolic biomarker signature associated with incident type-2 diabetes. A. Overall metabolic biomarker signature comparison of 135 metabolic biomarkers for prenatal famine exposure and incident type-2 diabetes (r = 0.77, p = 3×10^-27^) as established in the UK Biobank Study in a 12-year follow-up. **B.** Overall metabolic biomarker signature comparison of 135 metabolic biomarkers for prenatal famine exposure (excluding 101 individuals with type-2 diabetes from the analysis) and incident type-2 diabetes as established in the UK Biobank Study in a 12-year follow-up (r = 0.67, p = 1×10^-18^). The effect size estimates for each metabolic biomarker are shown as points. Famine-associated metabolic biomarkers are indicated in blue.

To explore whether the metabolic biomarker profile of prenatal famine exposure may reflect a risk of diseases beyond type-2 diabetes, we extended the analysis to recently published atlas of signatures of a wide range of diseases in the UK Biobank (4). We focused on metabolic biomarker signatures for the future onset of disease obtained from individuals not affected by the disease of interest at baseline. For the analysis, we utilized a subset of 154 common incident diseases that had at least one metabolic biomarker association in the UK Biobank (p < 5×10^-4^). Remarkably, the metabolic biomarker signature of prenatal famine exposure was positively correlated with the metabolic biomarker signature of 115 diseases (75%; 0.3 :< r :< 0.9, p < 3.2×10^-4^), and negatively correlated with 13 diseases (8%; -0.9 :< r :< -0.4, p < 3.2×10^-4^) (**Supplemental Table 4**). The metabolic biomarker signature of prenatal famine exposure exhibited the strongest correlation with the signature of the future risk of myocardial infarction (r = 0.9, p = 1.8×10^-47^). Other diseases with a strong correlation (r ≥ 0.7) included those related to the digestive system, diseases with an endocrine, nutritional and metabolic component, as well as diseases of the nervous system (**Fig. 3**). The diseases displaying a negative correlation were primarily associated with injury and other consequences of external causes, such as fractures and open wounds (**Supplemental Table 4**).

**Fig 3.**
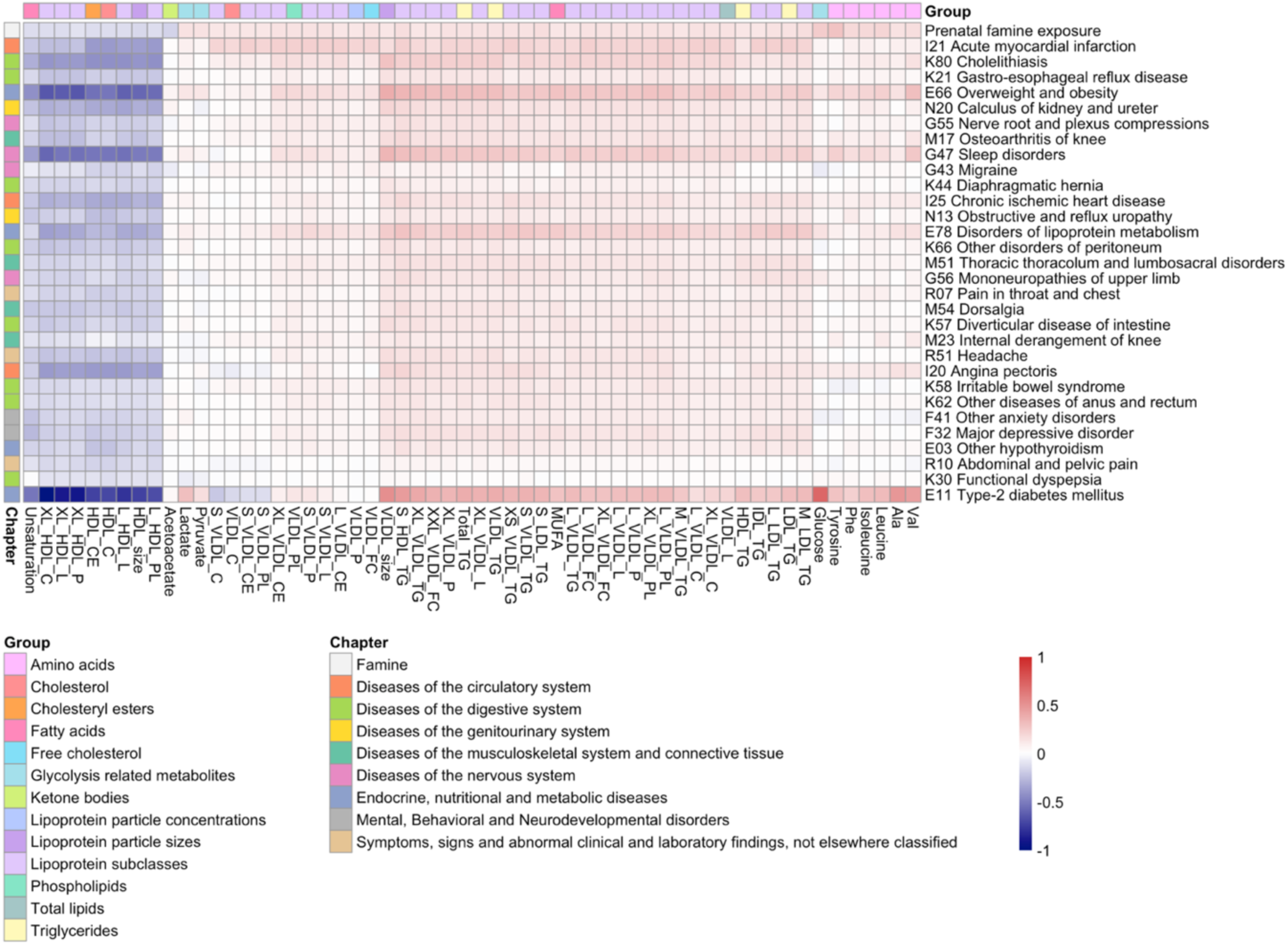
Correlation analysis of the metabolic biomarker signature associated with famine and the metabolic biomarker signature associated with various common diseases. Heatmap showing the effect size estimates of the 30 most correlated diseases to prenatal famine exposure. The columns are clustered according to the metabolic biomarker effect sizes and the rows are ordered according to the correlation of the metabolic biomarker signature of the disease to prenatal famine exposure (Pearson r for IK21 Acute myocardial infarction = 0.85, Pearson r for E11 Type-2 diabetes mellitus = 0.69). Only metabolic biomarkers that are nominally associated with prenatal famine exposure are shown (p < 0.05). The diseases are shown with their ICD-10 (International Classification of Diseases 10th Revision) classification. The full names of the metabolic biomarkers can be found in **Supplemental Table 2**.

We hypothesized that a potential common factor among the diseases with a similar metabolic biomarker signature is obesity. To test this hypothesis, we re-estimated the effect sizes for prenatal famine exposure while additionally adjusting for BMI and then re-calculated the correlation between the resulting BMI-adjusted metabolic biomarker signature with the signatures of the 154 common incident diseases (**Supplemental Table 4**). The strength of the correlations was consistently attenuated across all diseases (mean = -58%; SD = 19%). Among the 30 diseases whose metabolic biomarker signature was most similar to that of prenatal famine, the attenuation ranged between 27-48% and the correlations remained moderate (0.4 :< r :< 0.6). The degree of attenuation was not linked to whether the disease had an obvious metabolic component (**Fig. 4**).

**Fig 4.**
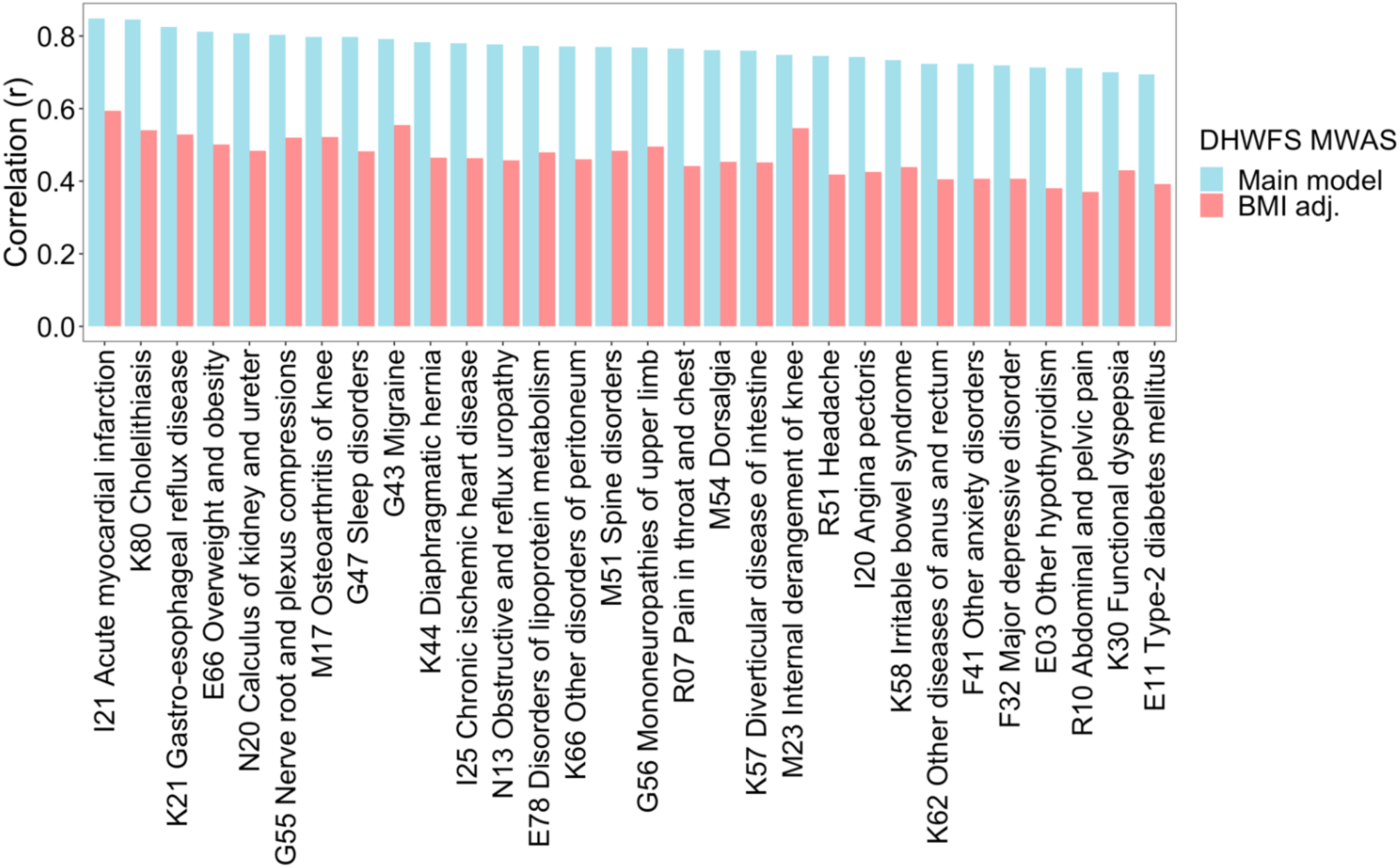
Effect of additional adjustment of BMI in the correlation analysis of the metabolic biomarker signature associated with famine and the metabolic biomarker signature associated with various common diseases. The main model within the DHWFS cohort was adjusted for age, sex, and cholesterol-lowering medication. The BMI-adjusted model within the DHWFS cohort was adjusted for age, sex, cholesterol-lowering medication, and BMI. The effect sizes estimated for these two models of prenatal famine exposure were each correlated to the effect sizes estimated for the risk of common diseases. The 30 diseases most correlated to the metabolic biomarker signature of prenatal famine exposure are shown.

## Discussion

We further defined the metabolic phenotype associated with prenatal famine exposure using nuclear magnetic resonance (NMR) metabolomic profiling. We show that prenatal exposure to undernutrition is associated with specific metabolic differences later in life, including higher levels of branched-chain amino acids (BCAA), an aromatic amino acid, and glucose. In addition, we report that the metabolic biomarker signature of prenatal famine has marked similarities to the signature of a wide range of common diseases.

Our study indicated specific differences in the metabolic profiles of famine-exposed individuals compared to controls. We observed that those exposed to famine prenatally have higher levels of the aromatic amino acid tyrosine, the branched-chain amino acids leucine and isoleucine, and glucose six decades after exposure. All four metabolic biomarkers have been linked to type-2 diabetes and thus support the known association between prenatal famine exposure and type-2 diabetes risk in adulthood (6,33). In addition, higher levels of other branched-chain and aromatic amino acids such as valine and phenylalanine that have also been associated with type-2 diabetes showed a nominally significant association with prenatal famine exposure, further supporting the link between famine and type-2 diabetes risk (33). Interestingly, the metabolic biomarker associations with prenatal famine were independent of BMI and type-2 diabetes status, indicating that they may not be fully driven by the higher BMI and increased prevalence of type-2 diabetes among famine-exposed individuals.

The link with a higher type-2 diabetes risk among individuals exposed to famine in the prenatal period was reinforced by investigating the complete range of metabolic biomarkers. We observed a strong resemblance in the metabolic biomarker signature of prenatal famine with that of the future onset of type-2 diabetes (33). Moreover, the strong correlation of the famine signature with the incident type-2 diabetes signature persisted after excluding participants who were already diagnosed with type-2 diabetes at the time of assessment. This result reinforces that type-2 diabetes is a main health outcome of prenatal famine exposure (6) and indicates that even exposed individuals not diagnosed with type-2 diabetes have an increased risk of developing this condition in the future.

Upon extending our analysis beyond type-2 diabetes, we observed a striking similarity between the metabolic biomarker signature of famine exposure and a wide range of other incident disease signatures. This included conditions like disorders of lipoprotein metabolism, obesity and type-2 diabetes, but also a priori less expected diseases like osteoarthritis, kidney stones, and depressive disorders. Interestingly, these high correlations were substantially attenuated for all incident diseases when we repeated the analysis using a metabolic biomarker profile of prenatal famine that was adjusted for BMI. Our findings suggest that BMI is a shared risk factor for or consequence of the diseases and that the metabolic biomarkers measured by the NMR platform used may have a particularly strong association with BMI. Of note, after accounting for BMI, moderate correlations between the metabolic biomarker profiles of incident disease and prenatal famine remained. Our findings and previous studies highlight that the NMR platform applied is especially useful for disease risk prediction, but of limited value to gain new mechanistic insights. Further studies with more comprehensive metabolomics platforms are needed to fully understand why the metabolic biomarker signature of prenatal famine exposure links to a broad range of diseases, including effects independent of obesity.

## Conclusions

Prenatal exposure to famine is associated with marked metabolic alterations later in life. Differences in individual metabolic biomarkers include higher levels of branched-chain amino acids, aromatic amino acids, and glucose. Moreover, the metabolic biomarker signature characteristic of prenatal famine strongly resembles that of a diverse set of diseases. Overall, our findings underscore the broad impact of prenatal famine on adult health and highlight obesity as a plausible contributing factor.

## Data Availability

The DHWFS data is available for replication purposes upon request to B. T. Heijmans and if replication is conducted within the secure Leiden University Medical Center network environment.

## Abbreviations

BCAA: branched-chain amino acids
BMI: body mass index
DHWFS: Dutch Hunger Winter Families study
GWAS: genome-wide association study
LMP: last menstrual period
NMR: nuclear magnetic resonance
SD: standard deviation

## Declarations

### Ethics approval and consent to participate

The Dutch Hunger Winter Families study was approved by the Medical Ethics Committee of Leiden University Medical Center (P02.082) and the participants provided verbal consent and written informed consent.

### Consent for publication

Not applicable.

### Availability of data and materials

The datasets supporting the conclusions of this article are included within the article and its additional files. The DHWFS data is available for replication purposes upon request to B. T. Heijmans and if replication is conducted within the secure Leiden University Medical Center network environment.

### Authors’ contributions

M.J.T., D.W.B., L.H.L. and B.T.H. were involved in the conception, design, and conduct of the study and interpretation of the results. M.J.T. performed the analyses and wrote the first draft of the manuscript, and all authors edited, reviewed, and approved the final version of the manuscript. B.T.H. is the guarantor of this work and, as such, had full access to all the data in the study and takes responsibility for the integrity of the data and the accuracy of the data analysis.

## Acknowledgements

Not applicable.

## References

1. Wishart DS. Metabolomics for Investigating Physiological and Pathophysiological Processes. Physiol Rev. 2019 Oct 1;99(4):1819–75.

2. Bragg F, Trichia E, Aguilar-Ramirez D, Bešević J, Lewington S, Emberson J. Predictive value of circulating NMR metabolic biomarkers for type 2 diabetes risk in the UK Biobank study. BMC Med. 2022 Dec 1;20(1).

3. Bell JA, Richardson TG, Wang Q, Sanderson E, Palmer T, Walker V, et al. Effects of general and central adiposity on circulating lipoprotein, lipid, and metabolite levels in UK Biobank: A multivariable Mendelian randomization study. The Lancet Regional Health - Europe [Internet]. 2022;21:100457. Available from: 10.1016/j.

4. Julkunen H, Cichońska A, Tiainen M, Koskela H, Nybo K, Mäkelä V, et al. Atlas of plasma NMR biomarkers for health and disease in 118,461 individuals from the UK Biobank. Nat Commun. 2023 Feb 3;14(1):604.

5. Guo Y, Chen S, Zhang Y, Wang H, Huang S, Chen S, et al. Circulating metabolites associated with incident myocardial infarction and stroke: A prospective cohort study of 90 438 participants. J Neurochem. 2022 Aug 15;162(4):371–84.

6. Lumey LH, Stein AD, Susser E. Prenatal famine and adult health. Annu Rev Public Health. 2011;32:237–62.

7. Liu H, Chen X, Shi T, Qu G, Zhao T, Xuan K, et al. Association of famine exposure with the risk of type 2 diabetes: A meta-analysis. Clinical Nutrition. 2020 Jun;39(6):1717–23.

8. Burger GCE, Drummond JC, Sandstead HR. Malnutrition and starvation in Western Netherlands: September 1944 - July 1945. General State Print. Office., The Hague; 1948.

9. Stein Z, Susser M, Saenger G, Marolla F. Famine and Human Development: The Dutch Hunger Winter of 1944-1945. New York: Oxford University Press; 1975.

10. Ravelli AC, van Der Meulen JH, Osmond C, Barker DJ, Bleker OP. Obesity at the age of 50 y in men and women exposed to famine prenatally. Am J Clin Nutr. 1999 Nov;70(5):811–6.

11. Stein AD, Kahn HS, Rundle A, Zybert PA, van der Pal-de Bruin K, Lumey LH. Anthropometric measures in middle age after exposure to famine during gestation: evidence from the Dutch famine. Am J Clin Nutr. 2007 Mar;85(3):869–76.

12. Lumey LH, Ekamper P, Bijwaard G, Conti G, van Poppel F. Overweight and obesity at age 19 after pre-natal famine exposure. Int J Obes (Lond). 2021 Aug;45(8):1668–76.

13. Ravelli GP, Stein ZA, Susser MW. Obesity in young men after famine exposure in utero and early infancy. N Engl J Med. 1976 Aug 12;295(7):349–53.

14. Lumey LH, Stein AD, Kahn HS, Romijn JA. Lipid profiles in middle-aged men and women after famine exposure during gestation: the Dutch Hunger Winter Families Study. Am J Clin Nutr. 2009 Jun;89(6):1737–43.

15. Ravelli A, van der Meulen J, Michels R, Osmond C, Barker D, Hales C, et al. Glucose tolerance in adults after prenatal exposure to famine. The Lancet. 1998 Jan;351(9097):173–7.

16. Lumey LH, Stein AD, Kahn H. Food restriction during gestation and impaired fasting glucose or glucose tolerance and type 2 diabetes mellitus in adulthood: Evidence from the DutchHunger Winter Families Study. J Dev Orig Health Dis. 2009 Apr;1:S164.

17. Tobi EW, Slieker RC, Luijk R, Dekkers KF, Stein AD, Xu KM, et al. DNA methylation as a mediator of the association between prenatal adversity and risk factors for metabolic disease in adulthood. Sci Adv. 2018 Jan;4(1):eaao4364.

18. Lumey LH, Stein AD, Kahn HS, van der Pal-de Bruin KM, Blauw GJ, Zybert PA, et al. Cohort profile: the Dutch Hunger Winter families study. Int J Epidemiol. 2007 Dec;36(6):1196–204.

19. WHO, IDF. Definition and diagnosis of diabetes mellitus and intermediate hyperglycaemia : report of a WHO/IDF consultation. World Health Organization; 2006. p.

20. Würtz P, Kangas AJ, Soininen P, Lawlor DA, Davey Smith G, Ala-Korpela M. Quantitative Serum Nuclear Magnetic Resonance Metabolomics in Large-Scale Epidemiology: A Primer on -Omic Technologies. Am J Epidemiol. 2017 Nov 1;186(9):1084–96.

21. Soininen P, Kangas AJ, Würtz P, Suna T, Ala-Korpela M. Quantitative serum nuclear magnetic resonance metabolomics in cardiovascular epidemiology and genetics. Circ Cardiovasc Genet. 2015 Feb;8(1):192–206.

22. 1000 Genomes Project Consortium, Auton A, Brooks LD, Durbin RM, Garrison EP, Kang HM, et al. A global reference for human genetic variation. Nature. 2015 Oct 1;526(7571):68–74.

23. Choi SW, O’Reilly PF. PRSice-2: Polygenic Risk Score software for biobank-scale data. Gigascience. 2019 Jul 1;8(7).

24. Smith CJ, Sinnott-Armstrong N, Cichońska A, Julkunen H, Fauman EB, Würtz P, et al. Integrative analysis of metabolite GWAS illuminates the molecular basis of pleiotropy and genetic correlation. Elife. 2022 Sep 1;11.

25. Homish GG, Edwards EP, Eiden RD, Leonard KE. Analyzing family data: A GEE approach for substance use researchers. Addictive behaviors. 2010 Jun;35(6):558–63.

26. Julkunen H, Cichońska A, Tiainen M, Koskela H, Nybo K, Mäkelä V, et al. Atlas of plasma NMR biomarkers for health and disease in 118,461 individuals from the UK Biobank. Nat Commun. 2023 Feb 3;14(1):604.

27. Tikkanen E, Jägerroos V, Holmes M V, Sattar N, Ala-Korpela M, Jousilahti P, et al. Metabolic Biomarker Discovery for Risk of Peripheral Artery Disease Compared With Coronary Artery Disease: Lipoprotein and Metabolite Profiling of 31 657 Individuals From 5 Prospective Cohorts. J Am Heart Assoc. 2021 Dec 7;10(23):e021995.

28. Al Rashid K, Taylor A, Lumsden MA, Goulding N, Lawlor DA, Nelson SM. Association of the functional ovarian reserve with serum metabolomic profiling by nuclear magnetic resonance spectroscopy: a cross-sectional study of ∼ 400 women. BMC Med. 2020 Aug 31;18(1):247.

29. Wang Q, Jokelainen J, Auvinen J, Puukka K, Keinänen-Kiukaanniemi S, Järvelin MR, et al. Insulin resistance and systemic metabolic changes in oral glucose tolerance test in 5340 individuals: an interventional study. BMC Med. 2019 Nov 29;17(1):217.

30. Santos Ferreira DL, Williams DM, Kangas AJ, Soininen P, Ala-Korpela M, Smith GD, et al. Association of pre-pregnancy body mass index with offspring metabolic profile: Analyses of 3 European prospective birth cohorts. PLoS Med. 2017 Aug;14(8):e1002376.

31. Tikkanen E, Jägerroos V, Holmes M V, Sattar N, Ala-Korpela M, Jousilahti P, et al. Metabolic Biomarker Discovery for Risk of Peripheral Artery Disease Compared With Coronary Artery Disease: Lipoprotein and Metabolite Profiling of 31 657 Individuals From 5 Prospective Cohorts. J Am Heart Assoc. 2021 Dec 7;10(23):e021995.

32. Guasch-Ferré M, Hruby A, Toledo E, Clish CB, Martínez-González MA, Salas-Salvadó J, et al. Metabolomics in Prediabetes and Diabetes: A Systematic Review and Meta-analysis. Diabetes Care. 2016 May;39(5):833–46.

33. Bragg F, Trichia E, Aguilar-Ramirez D, Bešević J, Lewington S, Emberson J. Predictive value of circulating NMR metabolic biomarkers for type 2 diabetes risk in the UK Biobank study. BMC Med. 2022 Dec 1;20(1).

