## Supplemental file for "Adults prenatally exposed to the Dutch Famine exhibit a metabolic signature associated with a broad spectrum of common diseases"

| Supplemental Table 1. Association of prenatal famine exposure with five metabolic biomarkers, as measured by NMR and clinical chemistry. | 2 |
| --- | --- |
| Supplemental Table 2. Association of prenatal famine exposure with 168 metabolic biomarkers. | 3 |
| Supplemental Table 3. Polygenic scores of metabolic biomarkers associated with prenatal famine exposure. | 4 |
| Supplemental Table 4. Correlation of the metabolic biomarker signature of prenatal famine exposure to the metabolic biomarker signature of diseases. | 5 |
| Supplemental Fig. 1. Flow chart of the study population. | 6 |
| Supplemental Fig. 2. Scatter plots and correlations between metabolic biomarkers measured by both NMR and clinical chemistry. | 7 |
| Supplemental Fig. 3. Clustering of the metabolic biomarkers according to their correlation. | 8 |
| Supplemental Fig. 4. Sex- and exposure timing-specific analyses on famine-associated metabolic biomarkers. | 9 |
| Supplemental information. BBMRI-NL Metabolomics consortium banner. | 10 |

**Supplemental Table 1. Association of prenatal famine exposure with five metabolic biomarkers, as measured by NMR and clinical chemistry.**

| Metabolic biomarker | n | Controls  Mean (SD) | Famine-exposed  Mean (SD) | Effect size (95% CI) | p value |
| --- | --- | --- | --- | --- | --- |
| *Triglycerides* |  |  |  |  |  |
| NMR | 928 | 1.57 (0.85) | 1.70 (0.95) | 0.15 (0.03, 0.27) | 0.012 |
| Clin. chemistry | 928 | 1.48 (0.85) | 1.62 (1.09) | 0.16 (0.04, 0.28) | 0.010 |
| *Total cholesterol* |  |  |  |  |  |
| NMR | 943 | 5.58 (0.99) | 5.61 (1.02) | 0.04 (-0.08, 0.16) | 0.501 |
| Clin. chemistry | 943 | 5.65 (1.04) | 5.72 (1.06) | 0.08 (-0.04, 0.19) | 0.202 |
| *HDL cholesterol* |  |  |  |  |  |
| NMR | 941 | 1.37 (0.35) | 1.32 (0.33) | -0.11(-0.22, 0.00) | 0.042 |
| Clin. chemistry | 941 | 1.58 (0.46) | 1.57 (0.45) | -0.02 (-0.13, 0.08) | 0.676 |
| *LDL cholesterol* |  |  |  |  |  |
| NMR | 918 | 2.34 (0.51) | 2.38 (0.51) | 0.08 (-0.05, 0.20) | 0.218 |
| Clin. chemistry | 918 | 3.41 (0.96) | 3.44 (0.93) | 0.04 (-0.08, 0.16) | 0.509 |
| *Glucose* |  |  |  |  |  |
| NMR | 938 | 5.22 (0.87) | 5.49 (1.18) | 0.24 (0.11, 0.36) | <0.001 |
| Clin. chemistry | 938 | 5.33 (1.00) | 5.55 (1.28) | 0.18 (0.06, 0.30) | 0.004 |

Clin. chemistry, clinical chemistry. CI, confidence interval. NMR, nuclear magnetic resonance. SD, standard deviation.

**Supplemental Table 2. Association of prenatal famine exposure with 168 metabolic biomarkers.**

See Excel file.

**Supplemental Table 3. Polygenic scores of metabolic biomarkers associated with prenatal famine exposure.**

| Metabolic biomarker | n independent SNPs ^a^ | Correlation coefficient (r) ^b^ | r^2 c^ | p value |
| --- | --- | --- | --- | --- |
| Tyrosine | 45 | 0.22 | 0.05 | 8.7x10^-12^ |
| Leucine | 16 | 0.11 | 0.01 | 6.0x10^-4^ |
| Glucose | 14 | 0.10 | 0.01 | 2.3x10^-3^ |

^a^Polygenic scores were computed using the independent GWAS hits at genome-wide significance (p value < 5x10^-8^) (Smith et al., 2022, eLife).

^b^Correlation coefficient for the correlation between the polygenic scores and the measured levels of the respective metabolic biomarker.

^c^Explained variance of the relationship between the polygenic scores and the measured levels of the respective metabolic biomarker.

**Supplemental Table 4. Correlation of the metabolic biomarker signature of prenatal famine exposure to the metabolic biomarker signature of diseases.**

See Excel file.

**n= 971**

Participants underwent clinical examination

**n= 9 excluded:**

No data on NMR metabolomics

**n= 962**

Participants with data on NMR metabolomics

**n= 944**

**Population for analysis:** fasted participants with data on NMR metabolomics

**n= 18 excluded:**

Non-fasted participants, n= 17

Metabolomics data outlier, n= 1

**n= 1075**

Participants completed the interview questions

**n= 165 excluded:**

No clinical examination

**Supplemental Fig. 1. Flow chart of the study population.**

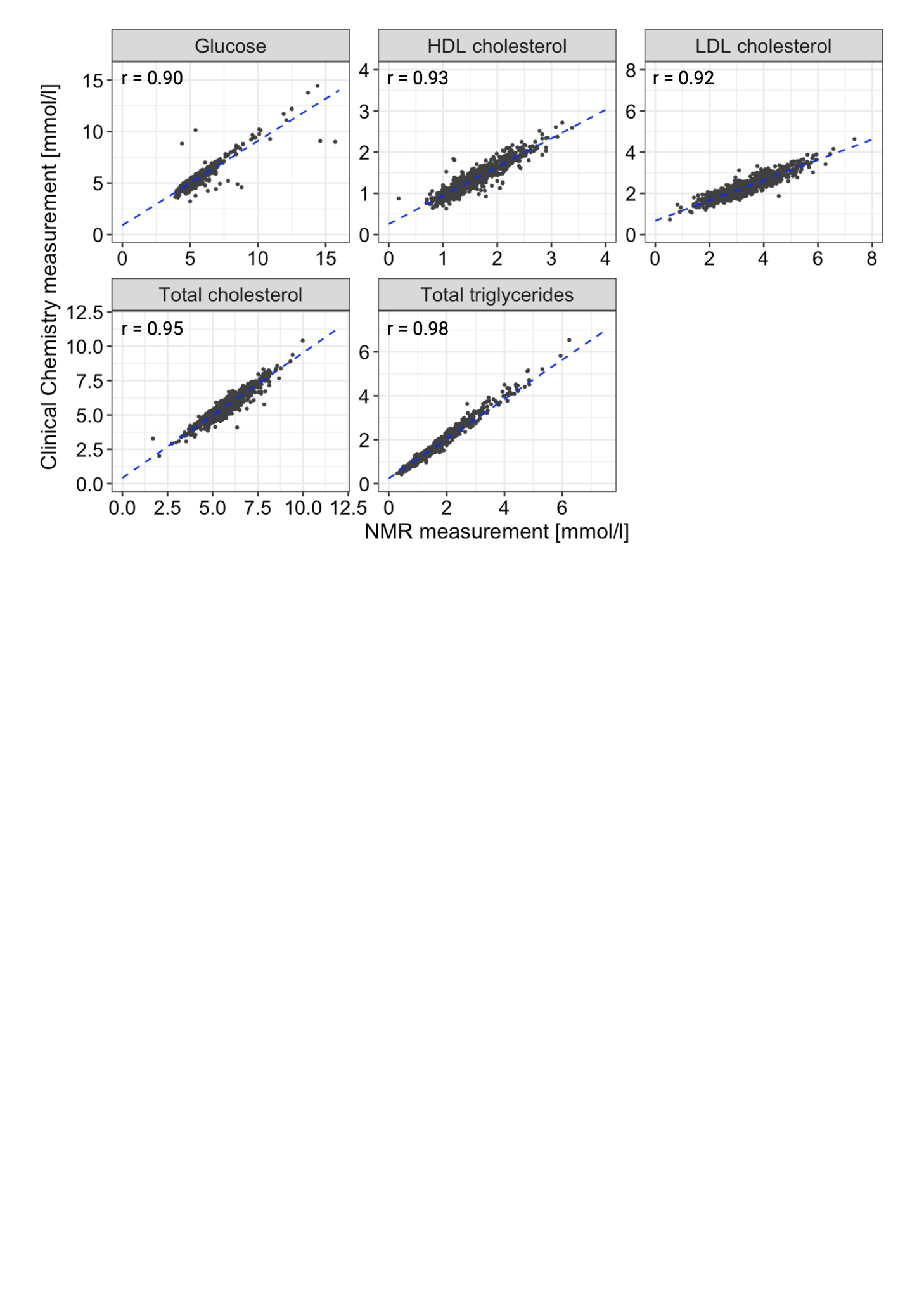

**Supplemental Fig. 2. Scatter plots and correlations between metabolic biomarkers measured by both NMR and clinical chemistry.** Metabolic markers were measured by both routine clinical chemistry and by an NMR metabolomics platform (Nightingale Health Ltd.) from serum samples. Pearson correlation coefficients (r) are depicted.

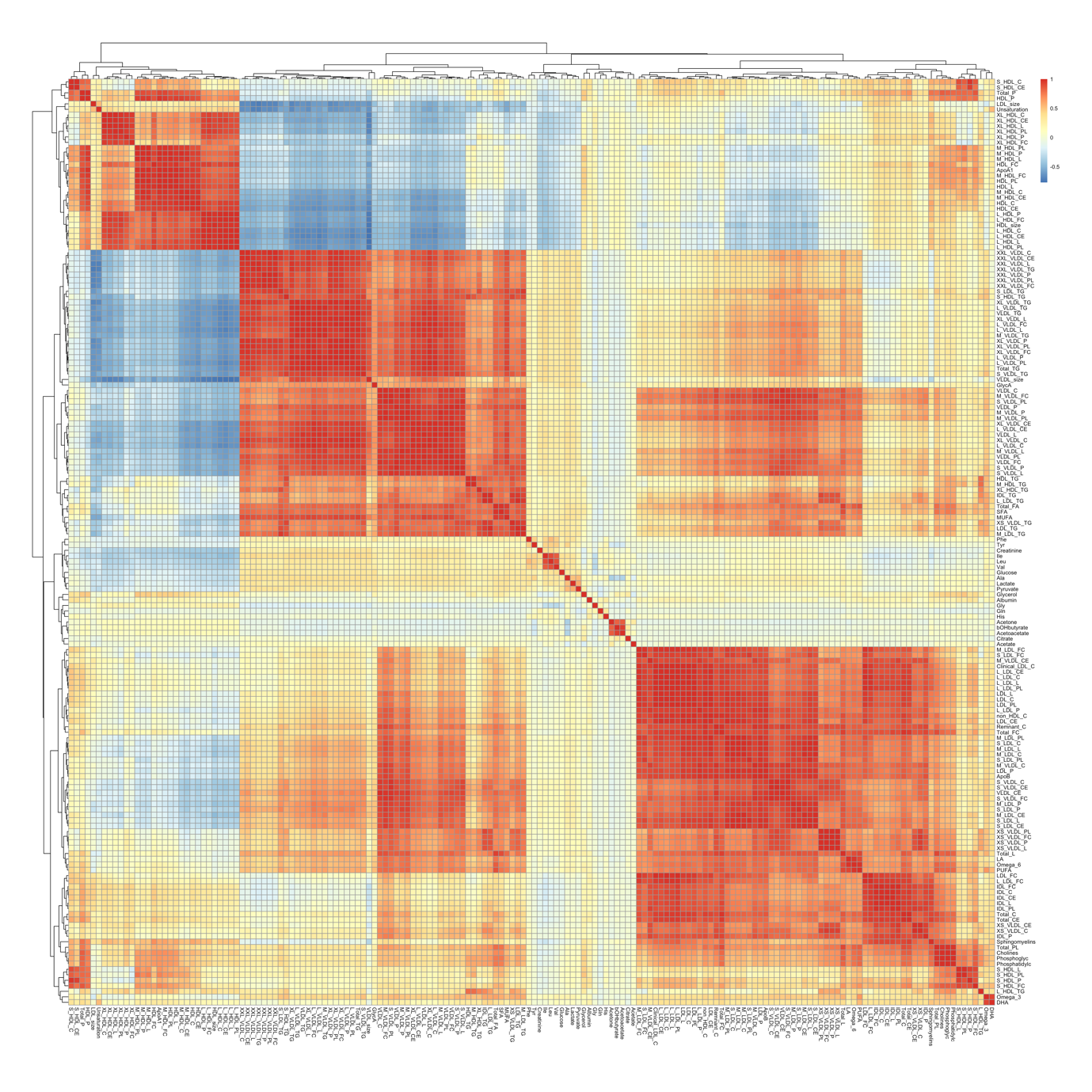

**Supplemental Fig. 3. Clustering of the metabolic biomarkers according to their correlation.** Heatmap showing the Pearson correlation coefficients among the 168 directly measured metabolic biomarkers included in the analysis. Red indicates high correlation, white indicates no correlation, and blue indicates high inverse correlation. The full list of metabolic biomarkers is found in **Supplemental Table 2**.

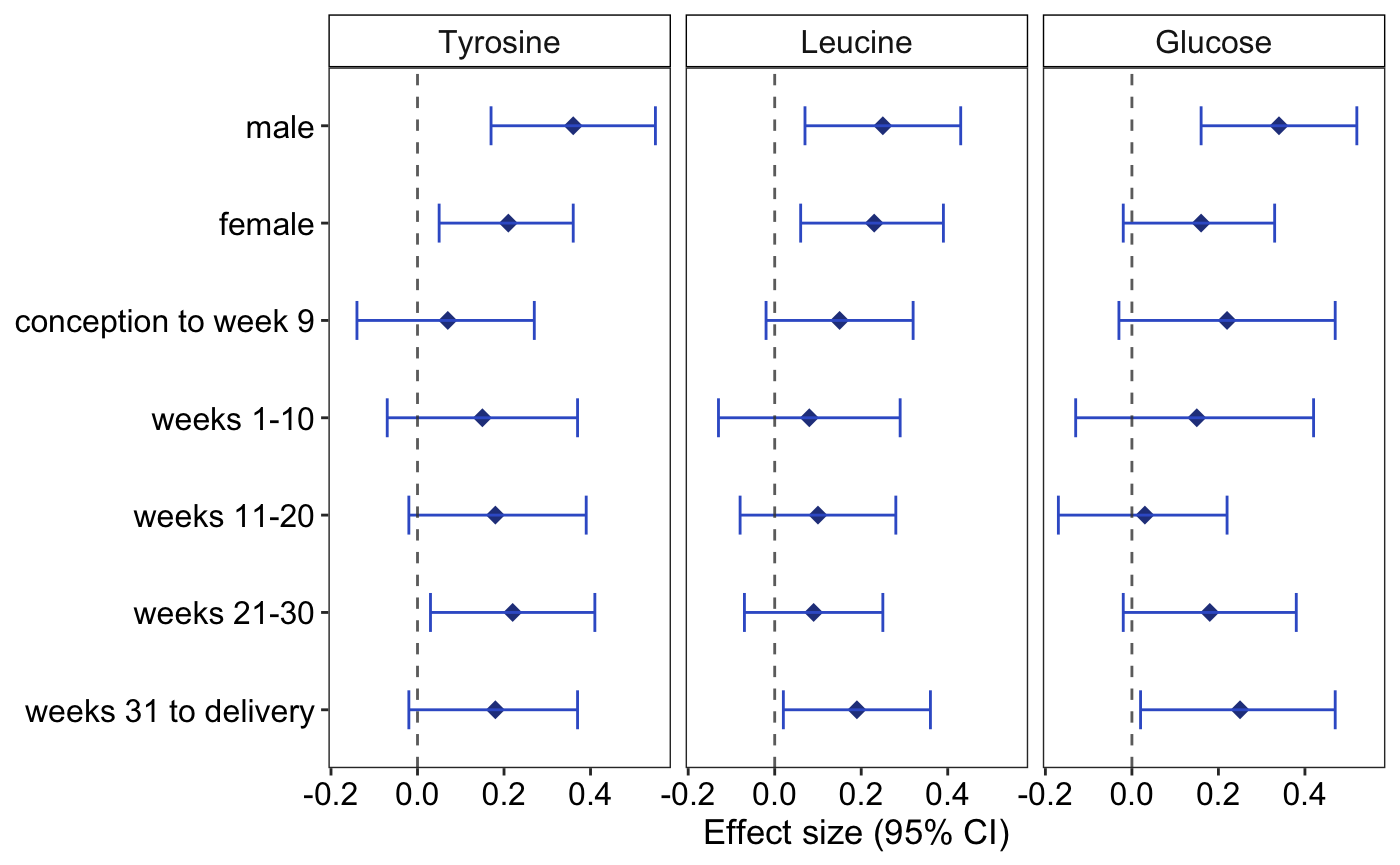

**Supplemental Fig. 4. Sex- and exposure timing-specific analyses on famine-associated metabolic biomarkers.** Sex-stratified analyses using regression adjusted for age and use of cholesterol-lowering drugs. Gestational timing-specific analyses were performed using linear regression in which the single indicator of famine exposure was replaced with indicator variables identifying exposure within each of the gestational time windows (exposed from conception to less than 10 gestation weeks, exposed gestation weeks 1 to 10, exposed gestation weeks 11 to 20, exposed gestation weeks 21 to 30 and exposed gestation weeks 31 to delivery), while adjusting for age, sex, and use of cholesterol-lowering drugs. Effect estimates and 95% confidence intervals are depicted for each model. Effect-size estimates are reported in standard-deviation (SD) units of the log-transformed metabolites.

**Supplemental information.** BBMRI-NL Metabolomics consortium banner.

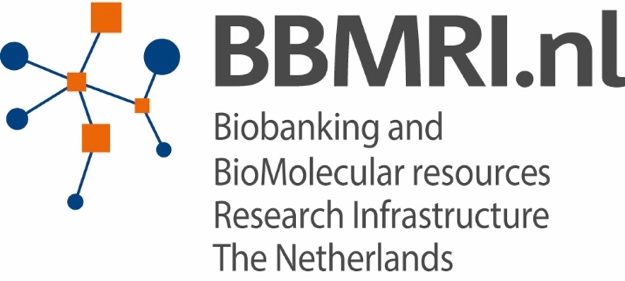
METABOLOMICS CONSORTIUM

Cohort Collection:

J.M. Geleijnse^1^, E. Boersma^2^, W.E. van Spil^3^, M.M.J. van Greevenbroek^4, 5^, C.D.A. Stehouwer^4, 5^, C.J.H. van der Kallen^4, 5^, I.C.W. Arts^5, 6, 7^, F. Rutters^8, 9^, J.W.J. Beulens^8, 9^, M. Muilwijk^8, 10^, P.J.M. Elders^8, 10^, L.M. 't Hart^8, 9, 11, 12^, M. Ghanbari^13, 14^, M.A. Ikram^13^, M.G. Netea^15^, M. Kloppenburg^16, 17^, Y.F.M. Ramos^18^, N. Bomer^19^, I. Meulenbelt^18^, K. Stronks^20^, M.B. Snijder^20^, A.H. Zwinderman^21^, B.T. Heijmans^18^, L.H.Lumey^22^, C. Wijmenga^23^, J. Fu^23, 24^, A. Zhernakova^23^, J. Deelen^25, 18^, S.P. Mooijaart^26^, M. Beekman^18^, P.E. Slagboom^18, 25^, G.L.J. Onderwater^27^, A.M.J.M. van den Maagdenberg^28, 27^, G.M. Terwindt^27^, C.Thesing^29, 8^, M. Bot^29, 8^, B.W.J.H. Penninx^29, 8^, S. Trompet^30, 26^, J.W. Jukema^30^, N. Sattar^31^, I.C.C. van der Horst^32^, P. van der Harst^33^, C. So-Osman^34, 35^, J.A. van Hilten^36^, R.G.H.H. Nelissen^37^, I.E. Höfer^38^, F.W. Asselbergs^39, 40^, P. Scheltens^41^, C.E. Teunissen^42^, W.M. van der Flier^43, 41^, J. van Dongen^29, 8^, R. Pool^29^, A.H.M. Willemsen^29, 8^, D.I. Boomsma^29, 8^

Sample Logistics, Database & Catalogue:

H.E.D. Suchiman^18^, J.J.H. Barkey Wolf^18^, M. Beekman^18^, D. Cats^45^, H. Mei^45^, M. Slofstra^23^, M. Swertz^46, 23^, M.J.T. Reinders^47, 48^, E.B. van den Akker^47, 18^

Steering committee:

D.I. Boomsma^29, 8^, M.A. Ikram^13^, P.E. Slagboom^18, 25^

Affiliations:

1. Division of Human Nutrition and Health, Wageningen University, Wageningen, The Netherlands
2. Thorax centre, Erasmus Medical Centre, Rotterdam, the Netherlands
3. Department of Rheumatology & Clinical Immunology, University Medical Center Utrecht, Utrecht, The Netherlands
4. Department of Internal Medicine, Maastricht University Medical Center (MUMC+), Maastricht, The Netherlands
5. School for Cardiovascular Diseases (CARIM), Maastricht University, Maastricht, the Netherlands
6. Department of Epidemiology, Maastricht University, Maastricht, the Netherlands
7. Maastricht Center for Systems Biology, Maastricht University, Maastricht, the Netherlands
8. Amsterdam Public Health Research Institute, Amsterdam, The Netherlands
9. Department of Epidemiology and Biostatistics, Amsterdam University Medical Center, Vrije Universiteit, Amsterdam, the Netherlands
10. Department of General Practice and Elderly Care Medicine, Amsterdam University Medical Center, Vrije Universiteit, Amsterdam, the Netherlands
11. Department of Epidemiology and Biostatistics, Amsterdam University Medical Center, Vrije Universiteit, Amsterdam, the Netherlands
12. Department of Cell and Chemical Biology, Leiden University Medical Center, Leiden, the Netherlands
13. Department of Epidemiology, Erasmus MC, University Medical Center, Rotterdam, The Netherlands
14. Department of Genetics, School of Medicine,, Mashhad University of Medical Sciences, Mashhad, Iran
15. Department of Internal Medicine and Radboud Center for Infectious Diseases, Radboud University Medical Center, Nijmegen, The Netherlands
16. Department of Clinical Epidemiology, Leiden University Medical Centre, Leiden, The Netherlands
17. Department of Rheumatology, Leiden University Medical Center, The Netherlands
18. Department of Biomedical Data Sciences, Section of Molecular Epidemiology, Leiden University Medical Center, Leiden, The Netherlands
19. Department of Experimental Cardiology, University of Groningen, University Medical Center Groningen, Groningen, The Netherlands
20. Department of Public Health, Academic Medical Center, University of Amsterdam, Amsterdam, The Netherlands
21. Department of Clinical Epidemiology, Biostatistics, and Bioinformatics, Academic Medical Centre, University of Amsterdam, Amsterdam, The Netherlands
22. Department of Epidemiology, Mailman School of Public Health, Columbia University, New York, NY 10032
23. Department of Genetics, University Medical Center Groningen, Groningen, The Netherlands
24. Department of Pediatrics, University Medical Center Groningen, Groningen, The Netherlands
25. Max Planck Institute for Biology of Ageing, Cologne, Germany
26. Department of Internal Medicine, Division of Gerontology and Geriatrics, Leiden University Medical Centre, Leiden, The Netherlands
27. Department of Neurology, Leiden University Medical Center, Leiden, The Netherlands
28. Department of Human Genetics, Leiden University Medical Center, Leiden, The Netherlands
29. Department of Biological Psychology, Amsterdam University Medical Center, Vrije Universiteit, Amsterdam, The Netherlands
30. Department of Cardiology, Leiden University Medical Center, Leiden, The Netherlands
31. Institute of Cardiovascular and Medical Sciences, Cardiovascular Research Centre, University of Glasgow, Glasgow, UK
32. Department of Critical Care, University Medical Center Groningen, Groningen, The Netherlands
33. Department of Cardiology, University Medical Center Utrecht, Utrecht, The Netherlands
34. Sanquin Blood Bank, Leiden and Department of Haematology, Groene Hart Hospital, Gouda, the Netherlands
35. International Society of Blood Transfusion (ISBT), Amsterdam, The Netherlands
36. Unit of Transfusion Medicine, Sanquin Blood Bank, Leiden, The Netherlands
37. Department of Orthopaedics, Leiden University Medical Center, Leiden, The Netherlands
38. Department of Clinical Chemistry and Hematology, UMC Utrecht, the Netherlands
39. Department of Cardiology, Division Heart and Lungs, University Medical Center Utrecht, Utrecht, The Netherlands
40. Julius Center for Health Sciences and Primary Care, University Medical Center Utrecht, Utrecht, The Netherlands
41. Department of Neurology & Alzheimer Center, VU University Medical Center, Amsterdam, The Netherlands
42. Neurochemistry Laboratory, Clinical Chemistry Department, Amsterdam University Medical Center, Amsterdam Neuroscience, The Netherlands
43. Department of Epidemiology and Biostatistics, VU University Medical Center, Amsterdam, The Netherlands
44. SURFsara, Amsterdam, the Netherlands
45. Sequence Analysis Support Core, Leiden University Medical Center, Leiden, the Netherlands
46. University of Groningen, University Medical Center Groningen, Genomics Coordination Center, Groningen, the Netherlands
47. Leiden Computational Biology Center, Leiden University Medical Center, Leiden, the Netherlands
48. The Delft Bioinformatics Lab, Delft University of Technology, Delft, the Netherlands
